# Predicted choice and acceptability of regimens for tuberculosis preventive treatment among people living with HIV in Uganda – a discrete choice experiment

**DOI:** 10.1101/2025.03.12.25323350

**Authors:** Hélène E. Aschmann, Allan Musinguzi, Jillian L. Kadota, Catherine Namale, Juliet Kakeeto, Jane Nakimuli, Lydia Akello, Fred Welishe, Anne Nakitende, Christopher Berger, David W. Dowdy, Adithya Cattamanchi, Fred C. Semitala, Andrew D. Kerkhoff

## Abstract

**Introduction:** Little is known about how people living with HIV would choose if offered different tuberculosis preventive treatment (TPT) regimens, and under which conditions they would accept treatment. Actionable evidence regarding preference for TPT is needed to inform policy and the development of novel TPT regimens.

**Methods:** Adults engaged in care at an HIV clinic in Kampala, Uganda, completed a discrete choice experiment survey with nine random choice tasks. In each task, participants first chose between two hypothetical TPT regimens with differing treatment features (number of pills, frequency, duration, adjusted antiretroviral dosage, and side effects). Second, they answered if they would accept the selected treatment, versus taking no treatment. We simulated predicted TPT regimen choice based on hierarchical Bayesian estimation of individual preference weights.

**Results:** Among 400 participants, 394 gave high-quality answers and were included (median age 44, 71.8% female, 91.4% previously received TPT). Across nine tasks, 60.2% (237/394) accepted all selected TPT regimens, 39.3% (155/394) accepted some regimens, and 0.5% (2/394) accepted none. Regimens requiring antiretroviral dosage adjustment were more likely to be unacceptable (adjusted odds ratio, aOR 27.4, 95% confidence interval [CI] 18.5 – 40.7), as were regimens requiring more pills per dose (aOR 24.5 [95% CI 16.6 – 36.3] for 10 pills compared to 1 or 5 pills per dose). Choice simulations showed that if only 6 months of daily isoniazid (6H) was available, 11.9% would prefer no TPT. However, offering a 4-pill, fixed-dose combination 3HP regimen in addition to 6H increased the acceptability from 88.1% to 98.8% (predicted choice of 3HP 94.5%, 6H 4.4%, no TPT 1.2%).

**Conclusions:** While adults living with HIV in Uganda demonstrate a high willingness to accept different TPT regimens, offering regimens with preferred features, such as 3HP as a fixed-dose combination, could drive TPT acceptance and uptake from high to nearly universal.

## Introduction

People living with HIV (PLHIV) in high tuberculosis (TB) incidence settings should receive TB preventive treatment (TPT).^1^ However, little is known under which circumstances people would prefer TPT or no TPT, as well as the specific regimens they would accept or reject. In 2024, 56% of PLHIV globally who newly initiated antiretroviral therapy (ART) received TPT.^2^ Offering short-course regimens as an alternative to six months of isoniazid (6H) is a key component of more person-centered TB prevention, aligning with the first pillar of the End TB Strategy.^3^ The World Health Organization (WHO) recommends several different short-course TPT regimens for PLHIV;^1^ however, the availability of these regimens differs by country.^4^

In Uganda, national guidelines recommend one of three TPT regimens for adult PLHIV: 6H, three months of weekly isoniazid and rifapentine (3HP), and one month of daily isoniazid and rifapentine (1HP).^5,6^ In practice, only 6H and 3HP are available. Despite achieving nearly 90% TPT coverage among PLHIV,^7^ person-centered strategies remain necessary to ensure high coverage among people newly enrolled in HIV care in Uganda.^2^ Furthermore, the duration of protection provided by TPT and the efficacy of repeat courses of TPT remains uncertain.^1,8,9^ Thus, understanding the acceptability of TPT regimens among all PLHIV in Uganda is crucial.

Although a few studies have described the preferences of children and their caregivers,^10,11^ evidence remains limited on the features and regimens influencing under what conditions PLHIV would accept TPT. Our prior work in Uganda showed that adult PLHIV preferred 3HP over 1HP,^12^ and highlighted that features beyond duration, including pill burden, dosing frequency, and the need for dosing adjustments to ART significantly influenced regimen choice.^13^ Similarly, for children, adolescents, and their caregivers, features like pill size, taste, and packaging have also been shown to be important.^11^ However, actionable evidence on expected choices of PLHIV between TPT or no TPT in the context of different regimen options, including those not yet introduced, is needed to guide policy and guideline recommendations.

Therefore, the objectives of this study were to 1) simulate the choice of TPT versus no TPT depending on the availability of different TPT regimens using data from a recent discrete choice experiment (DCE) among adult PLHIV in Uganda; 2) estimate the impact of different regimen features on TPT acceptability; and 3) evaluate associations between patient characteristics and a preference for no TPT.

## Methods

### Setting and participants

We conducted a cross-sectional DCE survey from July to November 2022, as previously described.^13^ The study took place at the Mulago Immune Suppression Syndrome (ISS) clinic, in Kampala, Uganda, the largest outpatient HIV clinic in the country. Briefly, individuals were eligible if they were enrolled in HIV care at the clinic, were 18 years or older, had not initiated a TPT regimen in the past year, and were not currently receiving TB treatment. We excluded individuals who did not provide informed consent or were currently in prison.

### Procedures

The survey was administered by trained interviewers on an electronic tablet. Before starting, participants were provided with general information on latent TB, and the DCE attributes and levels were explained with a structured flipbook, followed by a practice survey task.

### Ethics, consents, and permissions

The Makerere University School of Public Health Research and Ethics Committee, the University of California San Francisco Institutional Review Board and the Uganda National Council for Science and Technology approved the study. All participants provided written informed consent.

### DCE design

The DCE was designed in Sawtooth Lighthouse Studio version 9.13.2,^14^ with a list of attributes initially informed by the literature and iteratively refined through pilot testing. The final design included six attributes with 2 or 3 levels each: number of pills (1, 5, or 10 pills), frequency of dosing (weekly, twice per week, or daily), regimen duration (1, 3, or 6 months), adjusted HIV antiretroviral dosage (yes - second dose of ART needed, or no - no adjustment needed), mild side effects (10%, 50%, or 90% risk), and moderate or severe side effects (1%, 10%, or 20% risk) (**Figure S1**). Each participant completed nine random choice tasks and two fixed choice tasks: (1) a dominant choice task to assess comprehension of the exercise^15^ (**Table S1**), and (2) a task approximating 1HP and 3HP (**Table S2**). Choice tasks were unlabeled (Treatment “A” and “B”).

Participants first selected their preferred TPT regimen (A or B), and were then asked to choose between their preferred regimen and no TPT:

“If the TB prevention treatment you chose was available, would you actually take it? No treatment would mean:

- Your risk of developing TB is high. About 5 in 100 persons will get TB in the next three years.
- No additional tablets, just continue your usual ART.”

We based our estimate of TB incidence (5 in 100) among PLHIV on ART without TPT in Uganda on incidence rates reported in Uganda,^16^ a trial investigating early ART initiation without TPT,^17^ and the observation that TB incidence is about half as high for PLHIV on ART for longer than three years.^18^ Similar rates have since been reported based on registry data in Uganda.^19^ Participants with prior TPT experience were told research on repeat TPT is ongoing and “If … you were asked to take the therapy again, we are interested in understanding your preferences around the treatment regimen.”

As previously reported, we targeted a sample size of 400 participants to allow for robust subgroup analyses.^20^

### Statistical analysis

Simulations were performed using Lighthouse Studio version 9.13.2 (Sawtooth Software),^21^ and regressions were performed using R version 4.1.2.^22^ Prior to analysis, we conducted data cleaning to ensure the quality of individual-level respondent preference data. Participants with poor response quality – identified through incorrect answers to the dominant fixed choice task or other indications of inattention or incomprehension – were excluded from the analysis, as previously described.^13^ We analyzed which features made TPT regimens unacceptable in two ways: by mixed-effects logistic regression to account for individual-level variability,^23^ and by using choice simulations (described below).

For regression analyses, TPT regimens were categorized as either “acceptable” (i.e., the selected regimen was preferred over “no treatment”) or “unacceptable” (“no treatment” was preferred over the selected regimen), for each task. If no treatment was preferred over the selected regimen, both presented regimens in the task were classified as “unacceptable”. When a regimen was selected as preferred over the alternative regimen, and as preferred over no treatment, it was classified as “acceptable.” The alternative, less preferred regimen was not included in the regression, as it was unclear whether it would be preferred over no treatment. To minimize sparse data bias, variable categories (e.g., 1 and 5 pills) were grouped so that each combination was classified as acceptable or unacceptable at least 3 times. Side effects were only included in a sensitivity analysis for the same reason. Thus, the mixed effects logistic regression included four fixed effects (duration, frequency, number of pills per dose, and need to adjust ART medications, each with two categories) and a random intercept for individual participants.

For choice simulations, we used a hierarchical Bayesian model to calculate mean preference weights (i.e., part-worth utilities) for each attribute level. Using these preference weights, we applied a Shares of Preference Model using the Sawtooth Choice Simulator tool^24^ to predict participants’ choices. Parameters for existing regimens were defined as shown in **Table 1**. We simulated 1HP both with and without ART dosage adjustment required, as pharmacokinetic data suggest twice-daily dosing of dolutegravir, though definitive studies are lacking.^25^ Likelihoods of side effects were informed by previously reported adverse event rates.^26^ We performed choice simulations to: (1) predict preference for TPT over no TPT when only one attribute is changed at a time, using an ideal regimen and 3HP as references; (2) compare preferences for existing regimens; and (3) predict choices for the regimens historically available in Uganda. We selected 3HP as a reference regimen because this is increasingly becoming the standard TPT regimen in Uganda.^5^ Additionally, we simulated a scenario where 1HP was introduced alongside 6H and 3HP in the future.

**Table 1:** Simulation parameters of TPT regimens for choice predictions. 1HP: 1 month of daily isoniazid-rifapentine, 3HP: 3 months of weekly isoniazid-rifapentine, 3HR: 3 months of daily isoniazid-rifampin, 4R: 4 months of daily rifampin, 6H: 6 months of daily isoniazid, ART: Antiretroviral therapy, TPT: Tuberculosis preventive treatment

| **TPT** | **Duration [months]** | **Frequency** | **Pills per dose** | **Adjust ART dosage** | **Mild side effects [%]<sup>#</sup>** | **Moderate side effects [%]<sup>#</sup>** |
| --- | --- | --- | --- | --- | --- | --- |
| **1HP** | 1 | Daily | 4 | Yes* | 57.0 | 16.8 |
| **3HP** | 3 | Weekly | 4 | No | 11.5 | 6.0 |
| **3HR** | 3 | Daily | 4 | Yes | 29.7 | 2.3 |
| **4R** | 4 | Daily | 1 | Yes | 20.0 | 1.7 |
| **6H** | 6 | Daily | 2 | No | 36.1 | 8.2 |

\*As definitive studies are lacking, we also performed sensitivity analyses where 1HP was assumed to require no ART dosage adjustment. <sup>#</sup>Mild side effects were described as transient nausea, dizziness, fatigue, or skin rashes, for about a week, resolving on their own. Moderate or severe side effects were described as bothersome symptoms like tingling feet, allergy, or liver damage, requiring additional visits to the health care facility and which may require stopping medicine or being admitted to the hospital.

Finally, we performed a logistic regression to explore which patient characteristics were associated with refusing at least one TPT regimen across all random choice tasks. The baseline characteristics included as variables in the model were selected a priori based on the literature and the investigators’ hypotheses.

## Results

### Participant characteristics

We screened 456 persons, invited 414 to participate, and 401 provided consent. After excluding seven participants due to poor response quality (three failed the dominant task, three were likely inattentive, and one enrolled multiple times with inconsistent answers), we included 394 responses in the analyses. Most participants were female (71.8%), the median age was 44 years, and all were on ART, with a median duration of 10.4 years. Most had previously taken TPT (91.3%), either with 6H (66.7%), 3HP (32.5%) or both (0.8%) (**Table S3**).

### Predicted choices for existing regimens

When comparing the shares of preference for a choice between single existing regimens and no TPT, 3HR, 4R, and 1HP (if requiring ART adjustment) were unacceptable to a large proportion (range, 29-41%) of participants (**Figure 1A**). The standard 6H regimen was preferred over no TPT by 88.1%. The most acceptable regimens were 3HP as a fixed-dose combination (FDC) (4 pills; 99.1% preference over no TPT), and 1HP without ART adjustment (97.6% preference over no TPT).

**Figure 1:**
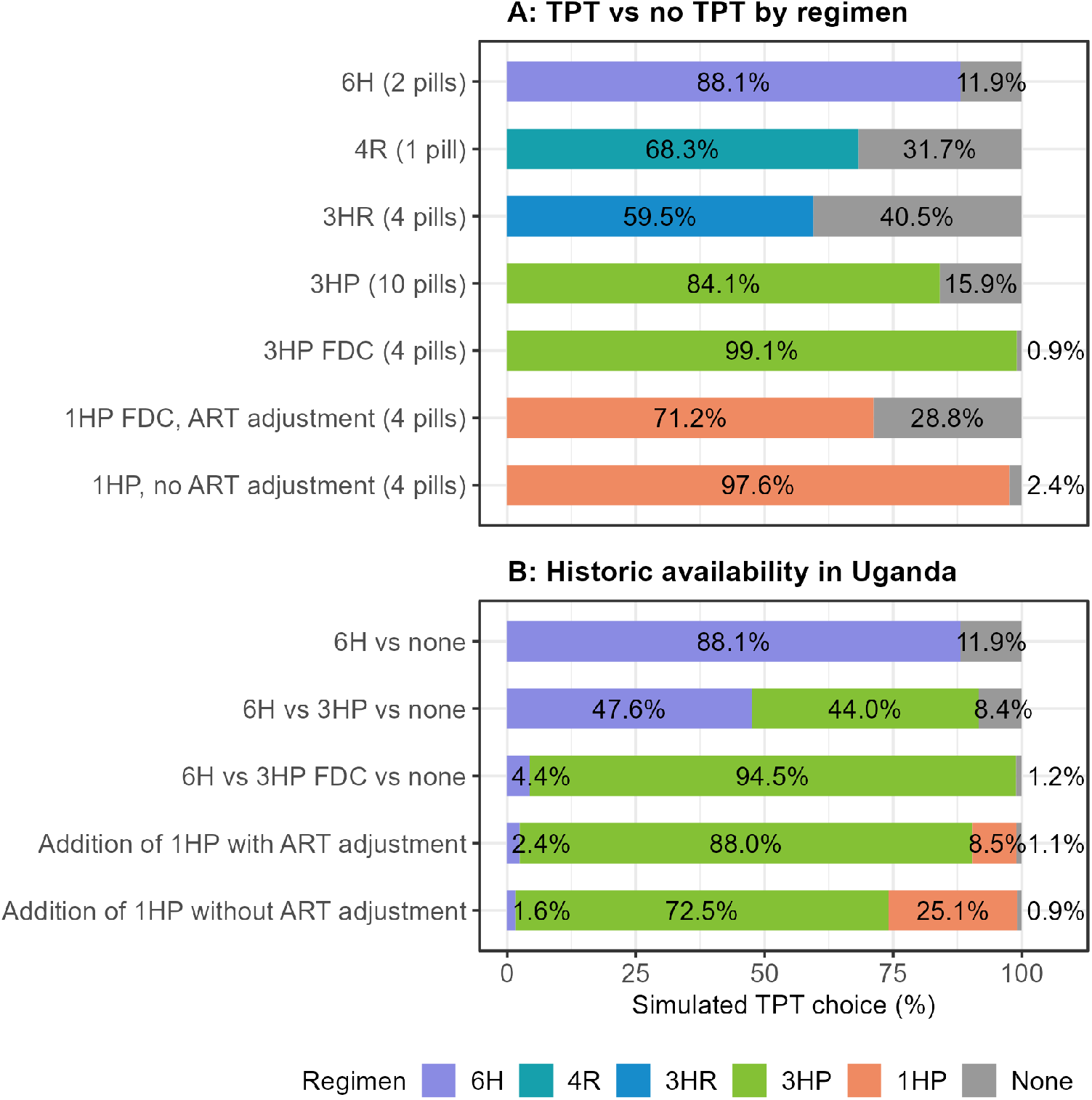
Choice simulations for existing TPT regimens. A) Acceptability of single regimens: choice simulation of TPT versus no TPT for all regimens recommended by the World Health Organization. B) Historically available regimens in Uganda: Until 2020, only 6H was available. 3HP was first available as a 10- or 11-pill regimen, then as a FDC with 4 pills. This simulation also includes a prediction of choices if 1HP were offered in the future. 1HP: 1 month of daily isoniazid and rifapentine, 3HP: 3 months of weekly isoniazid and rifapentine, 3HR: 3 months of daily isoniazid and rifampin, 4R: 4 months of daily rifampin, 6H: 6 months of daily isoniazid, ART: Antiretroviral therapy, FDC: fixed-dose combination, TPT: tuberculosis preventive therapy

In simulations of historically available options in Uganda, adding 3HP FDC to 6H reduced no-TPT preference to 1.2%, with 94.5% of participants predicted to choose 3HP (**Figure 1B**). In two scenarios, offering 1HP in addition to 3HP and 6H, showed little effect on the preference for no TPT. However, 25.1% would prefer 1HP if it required no ART adjustment.

We validated our choice simulations using the fixed choice task representing 3HP and 1HP. The observed preferences (85.0% 3HP, 14.2% 1HP, and 0.7% no TPT) were similar to the predicted probabilities (87.7%, 10.3%, and 2.0%, respectively). Predicted preferences for no TPT did not vary meaningfully by prior TPT experience (**Table S4** and **S5**).

### Impact of TPT features on acceptability

TPT regimens were substantially more likely to be considered unacceptable when they required 10 pills per dose compared to 1 or 5 pills (aOR 24.5 [95% CI 16.6 – 36.3]), ART dosage adjustment (aOR 27.4 [95% CI 18.5 – 40.7]), 3 or 6 months duration compared to 1 month (aOR 4.9 [95% CI 3.5 – 6.8]), and more frequent than weekly dosing (aOR 3.4 [95% CI 2.5 – 4.6]) (**Table 2**). In a sensitivity analysis, mild and moderate or severe side effects did not meaningfully influence the acceptability of a regimen and the results for other regimen features remained robust (**Table S6**). In a sensitivity analysis that distinguished all levels for duration, frequency, and number of pills, even regimens with 3-month compared to 1-month durations (aOR 4.0 [95% CI 2.7 – 5.9]), and 5 pills per dose compared to 1 pill per dose (aOR 4.7 [95% CI 3.1 – 7.1]) were more likely to be unacceptable; estimates for 10 vs 1 pill and ART dosage adjustment were unstable due to sparse observations of unacceptable regimens for some combinations (**Table S7**).

**Table 2:**
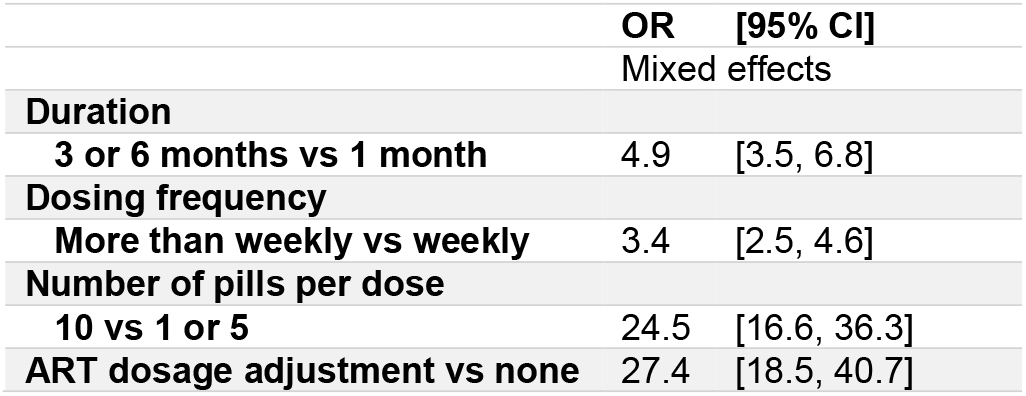
Regimen features associated with unacceptable vs acceptable TPT regimens. The analysis was conducted using mixed effects logistic regression.

|  | **OR** | **[95% CI]** |
| --- | --- | --- |
|  | Mixed effects |  |
| **Duration** |  |  |
| **3 or 6 months vs 1 month** | 4.9 | [3.5, 6.8] |
| **Dosing frequency** |  |  |
| **More than weekly vs weekly** | 3.4 | [2.5, 4.6] |
| **Number of pills per dose** |  |  |
| **10 vs 1 or 5** | 24.5 | [16.6, 36.3] |
| **ART dosage adjustment vs none** | 27.4 | [18.5, 40.7] |

*ART: antiretroviral therapy*

In simulations of an ideal TPT regimen (1 pill per dose, no ART adjustment, 1 month duration, weekly frequency, 10% risk of mild side effects, and 1% risk of moderate or severe side effects), 99.8% preferred TPT over no TPT (**Figure 2**). When modifying the ideal TPT regimen with one undesirable level at a time, 93.8% would still prefer a 10-pill-per-dose regimen over no TPT, and 96.2% would still prefer TPT if it required ART dosage adjustment. A preference for TPT over no TPT remained above 99% when the regimen required daily dosing or was associated with a high risk of side effects.

**Figure 2:**
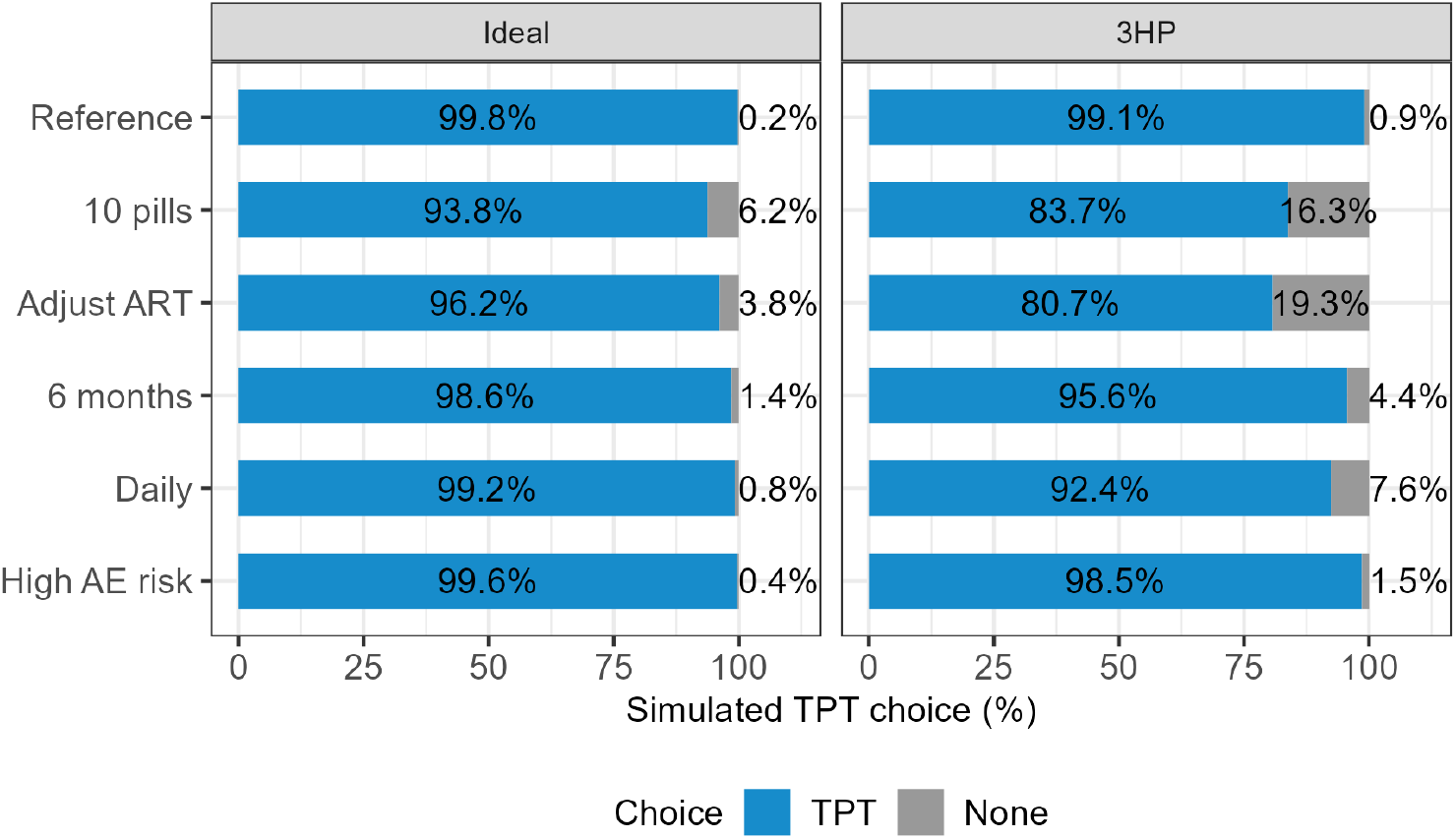
Predicted preference of regimens incorporating a single least preferred attribute level at a time in A) an ideal TPT regimen with otherwise optimal features (1 pill per week for 1 month, no ART adjustment, 10% risk of mild side effects, and 1% risk of moderate or severe side effects) and B) in a hypothetical TPT regimen that otherwise mirrors 3HP (4 pills per week for 3 months, no ART adjustment, 11.5% risk of mild side effects, and 6% risk of moderate or severe side effects).

In comparison, when 3HP was used as a reference it showed a 99.1% predicted preference over no TPT (**Figure 2**). If 3HP required ART dosage adjustment, 19.3% would prefer no TPT (18.4% increase in persons not accepting TPT). For a 10-pill 3HP regimen of 3HP, 16.3% would prefer no TPT (15.4% increase in persons not accepting TPT). 3HP was predicted to still be acceptable by more than 90% of participants if it required daily instead of weekly treatment, 6 months instead of 3 months duration, or had a high risk of side effects (90% for mild side effects, 20% for moderate or severe side effects).

### Individual variability in whether participants accept or refuse TPT regimens

Across 9 random choice tasks per participant, 60.2% (n=237/394) of participants accepted all regimens (i.e., preferred each chosen regimen over no TPT), 39.3% (n=155/394) accepted some chosen regimens, and 0.5% (n=2/394) accepted no chosen regimens. Any education (OR=5.3, 95% CI 2.8 – 10.9) and taking other medications (OR=2.4, 95% CI 1.5 – 3.9) increased the odds of refusing at least one chosen TPT regimen (**Table 3**). Longer time on ART decreased the odds of refusing any regimen (OR=0.72 per 5 years, 95% CI 0.54 – 0.97. Other participant characteristics, including sex, age, multidimensional poverty, employment status, prior TPT, prior active TB, and contraceptive use, were not associated with TPT regimen refusal.

**Table 3:** Association of participant characteristics with refusing any regimen versus accepting all regimens. The analysis was conducted using binary logistic regression.

|  | OR | [95% CI] |
| --- | --- | --- |
| **Female vs Male** | 1.03 | [0.61, 1.74] |
| **Age per 10 years (ref: 18)** | 1.24 | [0.94, 1.65] |
| **Education: any vs none** | 5.32 | [2.81, 10.9] |
| **Multidimensional poverty vs not\*:** | 1.34 | [0.75, 2.40] |
| **Working vs unemployed/other** | 1.14 | [0.65, 2.01] |
| **Prior TPT vs never** | 1.05 | [0.47, 2.39] |
| **Prior active TB vs none** | 1.39 | [0.77, 2.52] |
| **Years on ART (per 5 years, ref: 10.4)** | 0.72 | [0.54, 0.97] |
| **Contraceptive use vs none** | 1.67 | [0.84, 3.35] |
| **Other medications vs none** | 2.41 | [1.52, 3.85] |

\*The multidimensional poverty index captures deprivations in health, education, and living standards. We compared persons affected by multidimensional poverty or severe poverty versus persons vulnerable or not vulnerable to multidimensional poverty. ART: antiretroviral therapy, TB: tuberculosis, TPT: tuberculosis preventive therapy

## Discussion

In this simulation study using preference data from PLHIV in Uganda, we found that TPT with ART dosage adjustments and high pill burden was often unacceptable to PLHIV, particularly when paired with other less desirable regimen features like longer durations and daily dosing frequency. This suggests that minimizing treatment burden may be more critical for TPT acceptance than duration alone. 3HP as an FDC formulation emerged as the most preferred currently available TPT regimen, with 1HP showing high potential acceptability if ART dose adjustment proves unnecessary like for 3HP. 4R and 3HR, which are less utilized for adults across high burden settings, were predicted not to be acceptable to many participants. While the majority of PLHIV preferred all proposed regimens over no TPT, those with any education, shorter time on ART, and concurrent medications were more likely to refuse some TPT regimens.

Overall, participants were very willing to take TPT with preferable features, and in particular, 3HP as FDC or 1HP if it does not require ART dosage adjustments.^12^ While 1HP or 3HP are now available in more than 28 countries,^4^ there is limited published evidence about how their introduction has influenced TPT acceptance and uptake across different settings. In Pakistan, two recent studies reported 59% and 72% uptake of 3HP among household contacts in programmatic settings, respectively^27,28^, and although uptake was similar for 6H, completion rates were higher for 3HP.^28^ Early data from Ethiopia are encouraging, where the introduction of 3HP and its FDC formulation doubled TPT acceptance compared to 6H alone.^29^

Several factors likely influence the acceptance and completion of 3HP and other TPT regimens in real-world settings. A qualitative study in Uganda found that fear of contracting TB, awareness of being at risk of TB, and trust in healthcare workers facilitated acceptance and completion of 3HP.^30^ Another study in South Africa found that barriers to uptake are reduced after TPT education.^31^ Thus, the high acceptability in our study may reflect our educational approach using a pilot-tested education flipbook and our framing of TPT as a tradeoff against a 5% three-year TB risk, as well as familiarity of patients at the Mulago ISS clinic with TPT. A second qualitative study in our setting confirmed that shorter treatment duration and once-weekly dosing facilitated 3HP acceptance, while the high pill burden prior to FDC rollout was a barrier to completing treatment.^32^ In contrast to our finding that the risk of adverse events did not impact acceptability, this study also identified perceived 3HP safety as a facilitator for accepting 3HP.^32^ This may suggest that recommendations and shared positive experiences from peers and healthcare workers carry greater weight than being presented with varying risks of mild and moderate or severe side effects, which may be difficult for individuals to meaningfully interpret when deciding to start TPT. However, qualitative insights also indicate that experiencing side effects can lead to TPT discontinuation, particularly when symptoms interfere with daily activities and income generation.^32^

Person-centered preventive care is particularly important to promote uptake among persons who are likely to refuse offered regimens. We previously found that participants with fewer years of ART experience and those taking other medications more strongly preferred regimens that did not require ART dosage adjustment.^13^ Herein, we show that this also impacts the overall acceptability of TPT, with these groups being more likely to refuse TPT. Despite defining ART dosage adjustment as one additional pill of ART per day, its impact on the acceptability of TPT was much larger than for one additional pill per dose of TPT. Future research should explore and examine if alternative ways of explaining or framing ART dosage changes could mitigate some of the negative impacts on TPT acceptability.^33^ The higher refusal rate among more educated participants likely reflects an ability to evaluate different treatment options more critically, highlighting the need for patient-centered approaches and counseling that support informed decision-making across all education levels – particularly ensuring that those with lower health literacy have adequate support to make truly informed choices about TPT.

While offering a choice between 3HP and 1HP did not meaningfully reduce the proportion of participants declining all TPT options, this may reflect our highly treatment-experienced study population, social desirability bias, or other population-specific phenomena. Our findings showed that 25% of those who would accept 3HP would prefer 1HP (with no ART dosage adjustment) if available, demonstrating that providing TPT regimen choices better aligns with individuals’ preferences. Structured choice of 3HP and 1HP or other TPT alternatives that could become available in the future could have a greater impact on acceptability in more TPT-naive populations and in other settings. Recent findings from a large HIV prevention study in rural Uganda and Kenya demonstrate the potential of such approaches, where offering dynamic choice among post-exposure prophylaxis, oral, and long-acting injectable pre-exposure prophylaxis increased prevention coverage time from 13% to 70% and eliminated incident HIV cases.^34^ Similar approaches for TPT have yet to be evaluated, but they could increase population-level coverage by reaching people who might not otherwise accept available regimens.^35^

Key strengths of our study include that our choice simulations provide actionable evidence for and against the scale-up of different TPT regimens, the DCE methodology provides a robust approach to measuring preferences, and including an opt-out option after regimen selection enabled us to identify scenarios where even a “preferred” regimen was ultimately unacceptable – an insight that would be missed in forced-choice designs.^36–38^ Our study has some limitations. First, we could not validate participants’ hypothetical choices against actual treatment decisions. However, strong alignment between simulated predictions and a fixed choice task comparing 3HP versus 1HP provides some reassurance of response reliability and the predictive properties of the model.^39^ Second, despite special recruitment efforts to include TPT naïve persons, 91% of participants had prior TPT. We may have overestimated the acceptability of TPT (as most participants had prior TPT) or underestimated it (as we oversampled persons with no prior TPT). However, the main target audience for new TPT regimens may be people who would not accept current TPT options. Third, our results may not be generalizable beyond ART-experienced PLHIV in East Africa. Similar studies are needed among broader population groups and in other settings that also assess additional features that may be relevant to TPT decision-making beyond regimen choice. Finally, to avoid sparse data in our regression analysis, we had to combine some attribute levels, particularly grouping 1 and 5 pills versus 10 pills; we also did not include DCE levels for pill burden between 1 and 5, both of which limited our ability to detect more granular preferences between lower pill burdens.

## Conclusion

In conclusion, our choice simulations inform which existing TPT regimens should be prioritized to promote uptake and patient-centeredness among PLHIV in Uganda. 3HP as FDC, which is increasingly the standard regimen in Uganda and other high-burden countries, was highly acceptable to participants, and its continued rollout to replace 6H should be prioritized. Although offering 1HP alongside 3HP as FDC is not expected to substantially improve overall TPT uptake in our study population, it would enhance patient-centered preventive care by providing an option for those who prefer shorter duration over less frequent dosing, particularly if ART dosage adjustments prove unnecessary. More broadly, our results highlight the potential value of offering choices to increase population-level TPT coverage, and future research should explore the feasibility of providing and delivering such choices, especially as new TPT options with preferred features become available.

## Data Availability

De-identified individual participant data that support the findings of this study are available from the authors upon reasonable request after approval of a proposal.

## Acknowledgements

We thank the UCSF-Bay Area Center for AIDS Research (CFAR) for providing a Sawtooth license (P30 AI027763). We thank Barbara Alonso for creating beautiful icons for the levels in the discrete choice experiment.

This publication was supported by the National Center for Advancing Translational Sciences, National Institutes of Health, through UCSF-CTSI Grant Number UL1 TR001872. Its contents are solely the responsibility of the authors and do not necessarily represent the official views of the NIH.

