## Supplement 1 for "Predicted choice and acceptability of regimens for tuberculosis preventive treatment among people living with HIV in Uganda – a discrete choice experiment"

#### Tables

**Table S1**: Levels for dominant choice task

| | **Treatment A** | **Treatment B** |
|---|---|---|
| **Duration** | 6 months | 1 month |
| **Frequency** | Daily | Daily |
| **Number of pills** | 10 | 1 |
| **Adjust ART dosage** | No | No |
| **Mild side effects** | 90% | 10% |
| **Moderate side effects** | 20% | 1% |

ART: antiretroviral therapy

**Table S2**: Levels for fixed choice task representing 1HP versus 3HP

| | **Treatment A – 1HP** | **Treatment B – 3HP** |
|---|---|---|
| **Duration** | 1 month | 3 months |
| **Frequency** | Daily | Weekly |
| **Number of pills** | 5 | 5 |
| **Adjust ART dosage** | Yes | No |
| **Mild side effects** | 50% | 50% |
| **Moderate side effects** | 20% | 10% |

1HP: 1 month of daily isoniazid and rifapentine, 3HP: 3 months of weekly isoniazid and rifapentine, ART: antiretroviral therapy

**Table S3**: Participant characteristics at baseline, based on medical records and self-report

| Participants (N = 394) | N<br>median | (%)<br>(IQR) |
|---|---|---|
| **Female sex** | 283 | (71.8%) |
| **Age** | 44 | (IQR: 38, 51) |
| **BMI** | 26.2 | (IQR: 22.5, 30.1) |
| **Education** | | |
| None | 88 | (22.3%) |
| Primary | 148 | (37.6%) |
| Secondary | 115 | (29.2%) |
| Tertiary or higher | 43 | (10.9%) |
| **Work status** | | |
| Employed | 75 | (19.0%) |
| Self-employed | 240 | (60.9%) |
| Unemployed | 43 | (10.9%) |
| Not working | 22 | (5.6%) |
| Other | 14 | (3.6%) |
| **Multidimensional poverty index[1]** | | |
| Severely poor | 11 | (2.8%) |
| Poor | 59 | (15.0%) |
| Vulnerable | 110 | (27.9%) |
| Not vulnerable | 214 | (54.3%) |
| **Prior TPT** | | |
| None | 34 | (8.6%) |
| Prior 6H | 240 | (60.9%) |
| Prior 3HP | 117 | (29.7%) |
| Both prior 6H and 3HP | 3 | (0.8%) |
| **TPT completion (N=360)[2]** | | |
| TPT completed | 349 | (96.9%) |
| No, I discontinued LTBI treatment | 10 | (2.5%) |
| Do not know / do not want to answer | 1 | (0.3%) |
| **Experienced side effects from TPT (N=360)[2]** | | |
| Yes, and I had to see my doctor about it. | 28 | (7.8%) |
| Yes, but only mild ones and I did not see my doctor about it. | 57 | (15.8%) |
| No, I was fine. | 275 | (76.4%) |
| **History of active tuberculosis** | 71 | (18.0%) |
| **Current antiretroviral therapy** | | |
| Dolutegravir-based | 372 | (94.4%) |
| Efavirenz-based | 14 | (3.6%) |
| Other | 8 | (2.0%) |
| **Time on antiretroviral therapy (years)** | 10.4 | (IQR: 7.2,14.1) |
| **Viral load** | | |
| Suppressed | 388 | (98.5%) |
| Unsuppressed (≥1000 copies) | 3 | (0.8%) |
| Not yet done, recent HIV diagnosis | 1 | (0.3%) |
| Missing | 2 | (0.5%) |
| **Taking other medications[3]** | 145 | (36.8%) |
| **Herbal medicine use** | | |
| Within last month | 90 | (22.8%) |
| Within last year | 96 | (24.4%) |
| Longer than a year ago | 124 | (31.5%) |
| Never | 84 | (21.3%) |
| **Hormonal contraceptives among women (N=283)** | 52 | (18.4%) |

1. The multidimensional poverty index captures deprivations in health, education, and living standards.
2. Self-reported completion or side effects among those who reported prior TPT.
3. Currently taking other medications not including HIV medication or contraceptives.

3HP: 3 months of isoniazid and rifapentine, 6H: 6 months of isoniazid, BMI: body mass index, HIV: human immunodeficiency virus, IQR: interquartile range, TPT: tuberculosis preventive therapy.

**Table S4**: Simulated choice between 6H, 3HP, and no TPT by prior TPT experience. Participants with prior TPT experience were asked to imagine being offered repeat TPT, if it was proven effective.

| | Predicted choice 3HP | Predicted choice 6H | Predicted choice None |
|---|---|---|---|
| **All participants** | 94.5% | 4.4% | 1.2% |
| **No TPT experience** | 91.9% | 4.7% | 3.3% |
| **6H experience** | 93.6% | 5.1% | 1.3% |
| **3HP experience** | 96.3% | 3.6% | 0.2% |

3HP: 3 months of weekly isoniazid and rifapentine, 6H: 6 months of daily isoniazid, ART: Antiretroviral therapy, TPT: tuberculosis preventive therapy

**Table S5**: Simulated choice between 6H and no TPT by prior TPT experience. Participants with prior TPT experience were asked to imagine being offered repeat TPT, if it was proven effective. Thus, the interpretation is that 12.6% of participants with prior experience of 6H would not accept 6H if it was offered as a repeat regimen.

| | Predicted choice 6H | Predicted choice None |
|---|---|---|
| **All participants** | 88.1% | 11.9% |
| **No TPT experience** | 87.2% | 12.8% |
| **6H experience** | 87.4% | 12.6% |
| **3HP experience** | 90.1% | 9.9% |

3HP: 3 months of weekly isoniazid and rifapentine, 6H: 6 months of daily isoniazid, ART: Antiretroviral therapy, TPT: tuberculosis preventive therapy

**Table S6**: Sensitivity analysis for regimen features associated with unacceptable vs acceptable TPT regimens. This sensitivity analysis includes side effects, which were not included in the main analysis to address sparse data bias. The analysis was conducted using mixed effects logistic regression.

| | OR | [95% CI] |
|---|---|---|
| | Mixed effects | |
| **Duration** | | |
| **3 or 6 vs 1 month** | 4.9 | [3.5, 6.8] |
| **Dosing frequency** | | |
| **More than weekly vs weekly** | 3.4 | [2.5, 4.6] |
| **Number of pills per dose** | | |
| **10 vs 1 or 5** | 24.8 | [16.8, 36.8] |
| **ART dosage adjustment vs none** | 27.9 | [18.7, 41.4] |
| **Mild side effects** | | |
| **50% vs 10%** | 0.98 | [0.69, 1.40] |
| **90% vs 10%** | 0.90 | [0.64, 1.28] |
| **Moderate or severe side effects** | | |
| **10% vs 1%** | 1.24 | [0.87, 1.76] |
| **20% vs 1%** | 1.29 | [0.90, 1.83] |

ART: antiretroviral therapy, OR: odds ratio

**Table S7**: Sensitivity analysis for regimen features associated with unacceptable vs acceptable TPT regimens. This sensitivity analysis distinguishes all levels for duration, frequency, and number of pills per dose. Estimates for 10 pills per dose and ART dose adjustment are not robust and have wide confidence intervals. This is due to few observations of some combinations as unacceptable regimen, such as 1 weekly pill for 1 month. Estimates should therefore be interpreted with caution. The main analysis combines some levels to allow for robust estimates.

| | **OR** | **[95% CI]** |
|---|---|---|
| | Mixed effects | |
| **Duration** | | |
| **3 vs 1 month** | 4.0 | [2.7, 5.9] |
| **6 vs 1 month** | 11.6 | [7.4, 18.3] |
| **Dosing frequency** | | |
| **Twice per week vs weekly** | 1.8 | [1.2, 2.6] |
| **Daily vs weekly** | 10.5 | [6.8, 16.3] |
| **Number of pills per dose** | | |
| **5 vs 1** | 4.7 | [3.1, 7.1] |
| **10 vs 1** | 88.7 | [50.1, 157.0] |
| **ART dosage adjustment vs none** | 57.4 | [34.5, 95.5] |

### Figures

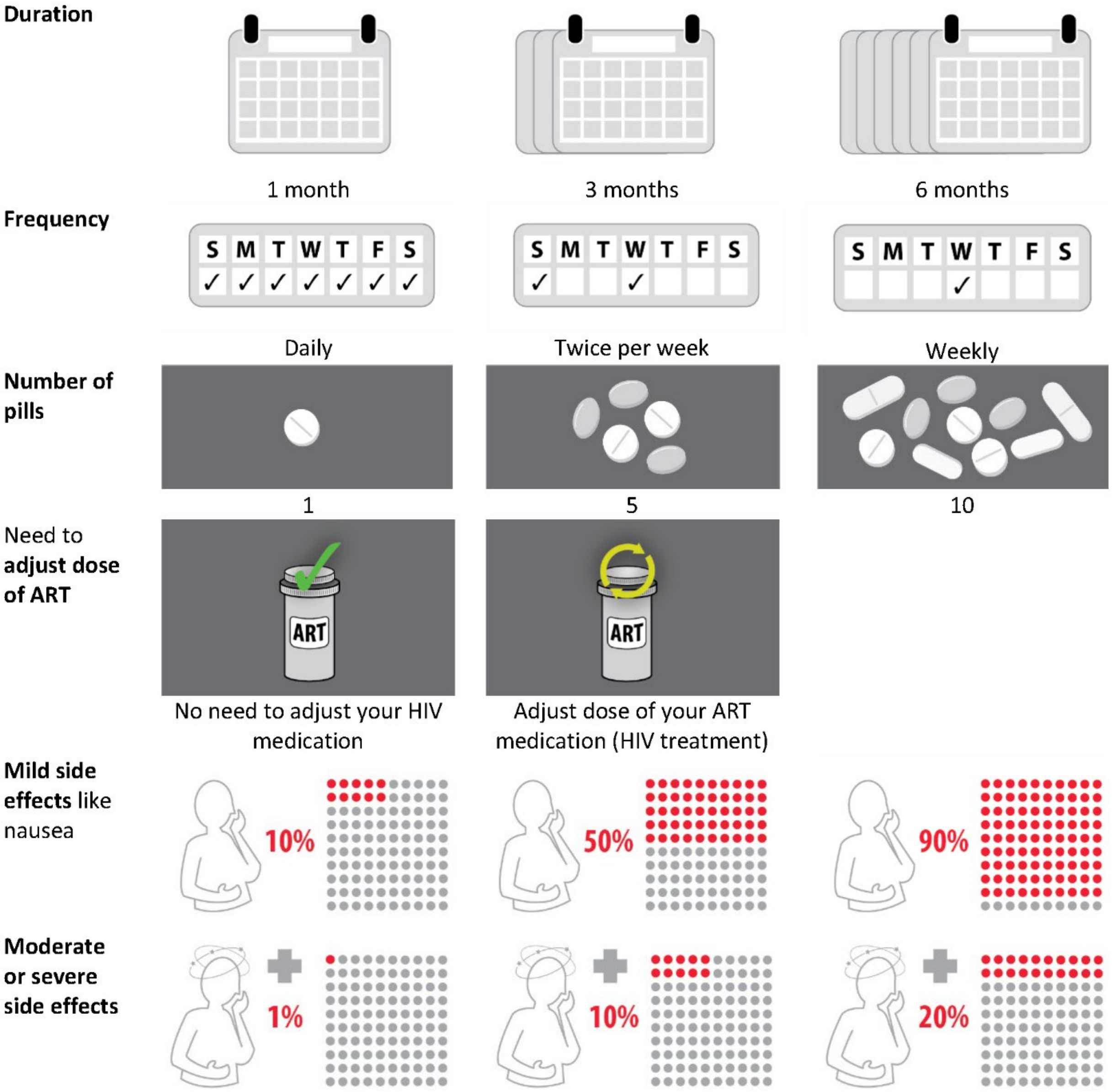


**Figure S1**: Attributes and levels in the discrete choice experiment describing different tuberculosis preventive treatment regimens. This figure shows the final selection of attributes (column 1) and how levels were depicted to participants (columns 2-4).
ART: antiretroviral therapy, HIV: human immunodeficiency virus

Reproduced from Aschmann, Hélène E., et al. "Preferences of people living with HIV for features of tuberculosis preventive treatment regimens in Uganda: a discrete choice experiment." Journal of the International AIDS Society 27.12 (2024): e26390.
